# Patient outcomes in a Type 1 hybrid implementation research on intravenous iron for anaemia in pregnancy in Nigeria: a mixed methods study

**DOI:** 10.1101/2025.05.05.25326718

**Authors:** Mobolanle Balogun, Opeyemi R. Akinajo, Rachel A. Thompson, Teniola Lawanson, Hameed Adelabu, Nadia A. Sam-Agudu, Bosede B. Afolabi

## Abstract

**Introduction:** Iron deficiency anaemia (IDA) is a common cause of anaemia in pregnancy (AIP), which is highly prevalent in low-and middle-income countries (LMICs). Evidence from LMICs indicates that intravenous (IV) iron formulations such as ferric carboxymaltose (FCM) are safe and effective alternatives to oral iron, and there is emerging data on implementation outcomes. Client outcomes are important additional considerations for successful scale-up of new interventions. We assessed patient experiences and satisfaction among women receiving IV versus oral iron for AIP in Nigeria.

**Methods:** This mixed-methods study was nested in a hybrid effectiveness-implementation trial that enrolled 1,056 pregnant women at 20-32 weeks gestational age. Participants were randomised to receive either FCM or oral ferrous sulphate (FS). Twenty-five percent of participants were sequentially sampled for interviewer-administered exit surveys and 66 women who received IV iron were purposively selected for in-depth interviews at last study visit. Quantitative data from the two treatment groups were evaluated using the Chi-squared test, while qualitative data were analysed thematically.

**Results:** We surveyed 252 participants; 128 treated with FCM and 124 with FS. Significantly higher proportions of the FCM (82.8%) vs FS group (63.7%) perceived their treatment to be “easy” (p = 0.006). Treatment was rated “excellent” by 80.5% of the FCM group compared to 62.1% of the FS group (p = 0.005). Women receiving FCM were more likely to recommend their treatment to a family member than the FS group (70.3% vs 50.0%, p= 0.018). The positive perception, experience and satisfaction with FCM was buttressed in the qualitative findings.

**Conclusion:** Positive patient experiences and satisfaction with IV iron positions this formulation as a viable alternative to oral iron for AIP in Nigeria. These patient-reported findings support available evidence on service and implementation outcomes and further support IV iron scale up in Nigeria and other African settings.

## Introduction

Anaemia in pregnancy (AIP) is a global health issue affecting both high and low-middle-income countries (LMICs) (1). The most common type of AIP is iron deficiency anaemia with an estimated prevalence of 50-75% worldwide (2,3). Evidence suggests that LMICs have significantly higher rates of AIP, possibly due to economic, sociological, and health factors related to pregnancy (4). In the African region, the estimated prevalence of AIP is about 43% (5). In Nigeria, about 20-40% of pregnant women are anaemic (6,7). AIP can lead to several adverse maternal and foetal outcomes, including increased risk of postpartum haemorrhage and intrauterine foetal death (8).

Oral iron is the standard treatment in Nigeria for mild to moderate cases of AIP (9). However, the most used form of oral iron, ferrous sulphate (FS), is linked with various side effects and low adherence to treatment (7,10). Additionally, due to poor health-seeking behaviour and financial constraints, many women fail to complete the standard oral iron regimen (11). This non-compliance with oral iron increases the risk of undertreated and persistent AIP in pregnancy (3,12). Intravenous (IV) iron has proven to be a viable alternative to oral iron for the treatment of AIP (13). Previously, high-molecular-weight IV iron formulations were infrequently used in clinical practice due to their tendency to cause severe allergic reactions and anaphylaxis (7,13). However, newer third-generation, low-molecular-weight formulations—like ferric carboxymaltose (FCM)—are both effective and safe for use during pregnancy (14).

FCM is a recent iron formulation devoid of dextran, featuring a nearly neutral pH, physiological osmolarity, and enhanced bioavailability (15). This formulation enables a single-dose administration with a 15-20 minute infusion time and the option for higher dosages (up to 1,000 mg) (14,15). These characteristics make FCM a compelling alternative for other iron formulations, considering factors such as risk profile, effectiveness, patient comfort, convenience, and the utilisation of staff and institutional resources (14,15).

Several studies indicate that FCM is faster and safer than oral iron therapy for treating AIP, and similar trials have been conducted in other LMICs, and are advocating for, or using FCM for treating AIP (13,14). It is essential to acknowledge that demonstrating the effectiveness of an intervention, such as FCM, does not automatically equate to its widespread acceptance and use. Therefore, while determining the effectiveness of this treatment is vital, the success of its implementation is equally significant. One key factor in understanding the context for implementation success is a focus on user satisfaction, which is crucial to how well this intervention is embraced and integrated within a healthcare system (16). Since FCM is a new treatment in the Nigerian context, it is imperative to assess users’ perceptions and satisfaction levels with IV iron compared to oral iron. Comparisons of treatment perceptions and experience between women receiving FCM and standard of care (FS) would help to better understand and address the preferences of AIP patients, given that experience of care is an important indicator of quality (17).

Thus, the objectives of this study were to assess and evaluate patient satisfaction, compare the perceptions of women who received IV versus oral iron for the treatment of AIP, and gain a deeper understanding of their experiences and the factors that influence the uptake of this intervention.

## Methods

### Study design

This mixed-methods study employs a convergent parallel design, combining quantitative and qualitative data collection methods to provide a comprehensive perspective on the users’ perceptions, experience and satisfaction with IV iron therapy. This approach allows for an exploration of the relationship between the two methodologies, offering a deeper understanding of the research topic and examining how the results from each method support or contradict one another (18). This study was conducted as a part of a hybrid type 1 effectiveness-implementation study that compared IV and oral iron in the treatment of iron deficiency anaemia in pregnant women in Nigeria (IVON) (6). As depicted using the Proctor’s framework for implementation outcomes, the implementation outcomes assessed in the trial were acceptability, feasibility, fidelity and cost-effectiveness; and the service outcomes assessed were effectiveness and safety (6,11). This current study assesses patient (client) outcome of satisfaction (Fig 1). To present our findings, we followed the Standards for Reporting Implementation Studies (StaRI) checklist to report our findings (S1 Checklist) (19).

**Fig 1.**
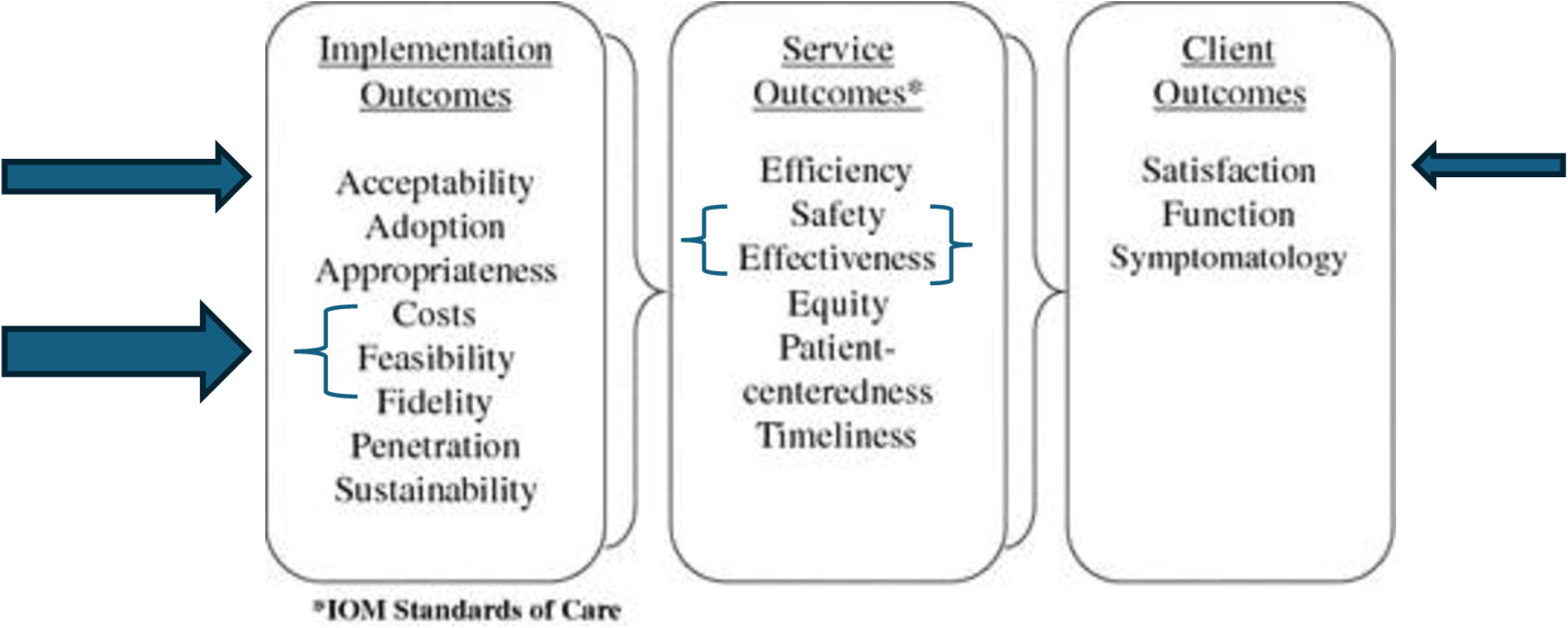
Representation of Proctor’s 2011 schematic with Implementation, Service and Client Outcomes relevant to the IVON Trial.

### Study setting

This study was conducted in Nigeria, with a population of over 200 million people, an average of seven million births annually, and which accounts for 29% of all maternal deaths worldwide (20,21). Selected study states were the two most populous states in the country: Kano in the North-West and Lagos in the South-West (22). The two states have different cultures, educational levels, and healthcare service utilization (22). The 11 public health facilities involved in the IVON trial included two primary health centres (PHCs) in Lagos, three PHCs in Kano, and two secondary and one tertiary facility in each state. The IVON trial randomly assigned eligible pregnant women between 15 and 49 years old and 20-32 weeks gestational age into the oral FS (control) arm or IV FCM (intervention) arm. A total of 1,056 women participated in the trial, with 527 in the FCM group and 529 in the oral group, all of whom had provided written informed consent.

### Participants recruitment and sampling technique

#### Quantitative phase

The study involved postpartum women who had received FCM or FS between 20-32 weeks gestational age who were followed up and monitored from enrollment, with regular checks every four weeks until 28 weeks, biweekly until 36 weeks, and weekly until delivery. Additionally, these women were followed up at two-week and six-week postpartum clinic visits following delivery. Participants were eligible for an exit survey at their scheduled postnatal clinic visit at six weeks postpartum, the timepoint at which their follow-up ended in the IVON trial, and participants completed and exited the trial. We aimed to survey 25% of women from each study arm, which equated to every fourth participant, thereby targeting a total of 132 participants in each group and 264 in total. The contribution of each facility was adjusted based on the total number of participants enrolled.

#### Qualitative phase

We purposively selected participants who had IV iron therapy based on various characteristics such as age, parity, prior experience with oral iron therapy and those who receive care from healthcare facilities known for their high patient volumes. We also considered unique circumstances surrounding the administration of IV iron therapy such as the length of waiting time for receiving IV iron, any apprehensions such as needle pricks, heightened anxiety related to the administration process and occurrence of side effects if any. We used the concept of information power to ensure that our gathered data was both sufficient and relevant (23). This approach considers the study’s specific objectives, the unique characteristics of the sample, and the valuable, high-quality insights provided by the users, allowing us to gain a comprehensive understanding of their experiences with the intervention (23).

#### Data collection tools

The research team designed a 10-item survey tool to collect data on participant’s perceptions and satisfaction regarding the AIP treatment (FCM or FS) they received during the trial. The survey assessed trial participants’ perceptions about the communication from providers about their treatment; the challenges experienced during the treatment, including side effects; overall satisfaction with their treatment; and recommendation of treatment to a family member. The questions had Likert scale responses that were relevant for each question e.g., never, a few times, most of the time and all the time; and excellent, good, and fair. Before data collection, the survey underwent a rigorous review process, was corrected by the research team, and was piloted with non-participating pregnant women to ensure its validity and reliability.

A semi-structured topic guide with five sections was developed to explore users’ knowledge of AIP, their perceptions of and relative advantages of IV iron compared to oral iron, and their challenges and experiences with the administration process, including overall satisfaction with IV iron therapy. This guide was used for qualitative data collection through in-depth interviews (IDIs) with participants selected purposively.

#### Data collection methods

Participants were contacted before their six-week postpartum visit to inform them about the exit survey and the IDIs and to ascertain their interest in participating. Of the 264 participants contacted across all the healthcare facilities, 260 were reachable, and 252 agreed to participate in the exit surveys. On the day of their postpartum visit, participants who agreed to participate were interviewed after informed consent was obtained. Eight trained research assistants who were independent of the clinical trial conducted the interviewer-administered surveys. The 30–45-minute surveys were administered in a private room at the health facility after each participant was discharged from the trial by the research staff; these were efforts to reduce social-desirability bias.

For the IDIs, we piloted the interview guide before data collection and made necessary adjustments based on the participants’ responses. We chose IDIs as they provided a practical opportunity for one-on-one discussions with the participants who received IV iron therapy (24). In total, we interviewed 66 participants, purposively selected from all eleven healthcare facilities. All interviews were conducted by the same trained research assistants who administered the surveys in private rooms at the healthcare facilities. Each interview lasted between 15 and 40 minutes. We repeated participants’ responses as needed for accuracy and clarity of intended meanings.

All participants were compensated for their time with transportation fare. Data were collected between October 2021 and January 2023. Quantitative data were collected using an electronic device (tablet) and entered directly into Research Electronic Data Capture (REDCap) (25,26). For the qualitative data, we obtained verbal consent from each participant before commencing the audio recordings. Subsequently, each interview was audio-recorded, and all interviews were transcribed verbatim.

## Data analysis

### Quantitative data analysis

STATA version SE15.1 (StataCorp, College Station, Texas, USA) was used to analyze the data. The continuous variable, age, was tested for normality using the Shapiro-Wilk test. Descriptive statistics, such as frequencies, percentages, median and interquartile range, were used to describe the sociodemographic characteristics of the participants. Differences in categorical variables between the two groups were evaluated using the Chi-squared test. The level of significance was set at 0.05.

### Qualitative data analysis

All transcribed interviews were reviewed for accuracy and completeness and analyzed using rapid thematic analysis. Two qualitatively trained researchers coded a subset of the transcript independently using a deductive approach to create a codebook, which was applied to the rest of the dataset. Next, we developed, reviewed, and refined our themes and sub-themes to accurately reflect the dataset’s meaning. The analysis was conducted manually.

### Integration of Quantitative and Qualitative Data

In this study, we utilized two complementary data collection methods, each holding equal significance in our research design. To effectively integrate the findings from these datasets, we employed a joint display matrix which, enabled us to systematically compare and contrast the varied findings, highlighting the relationships between the numerical data and the richer contextual information derived from the qualitative insights (27).

### Ethics

Ethical approval was obtained from the National Health Research Ethics Committee of Nigeria (NHREC/01/01/2007-17/01/2021), Health Research and Ethics Committees of the Lagos University Teaching Hospital (ADM/DCST/HREC/APP/3971), Aminu Kano Teaching Hospital, Kano State (NHREC/28/01/2020/AKTH/EC/2955) and Ministry of Health, Kano State (MOH/Off/797/T.1/2102). Social approval was obtained from the Lagos State Health Service Commissions (LSHSC/2222/VOLIII) and the Lagos State Primary Health Care Board (LS/PHCB/MS/1128/VOL.VII/100) for using the sites in Lagos state. As this study involved minimal risk of harm to participants, we obtained verbal informed consent from all participants before data collection.

## Results

### Quantitative results

A total of 252 women participated in the survey; 128 had been treated with FCM while 124 had been treated with FS. Median age was 27 years (IQR: 22-32) with age ranged between 16 and 44 years. Most participants were married (96.2%), attended secondary health facilities, and resided in urban areas (94%); 49.6% had received secondary education and 44.4% were self-employed (Table 1).

**Table 1:**
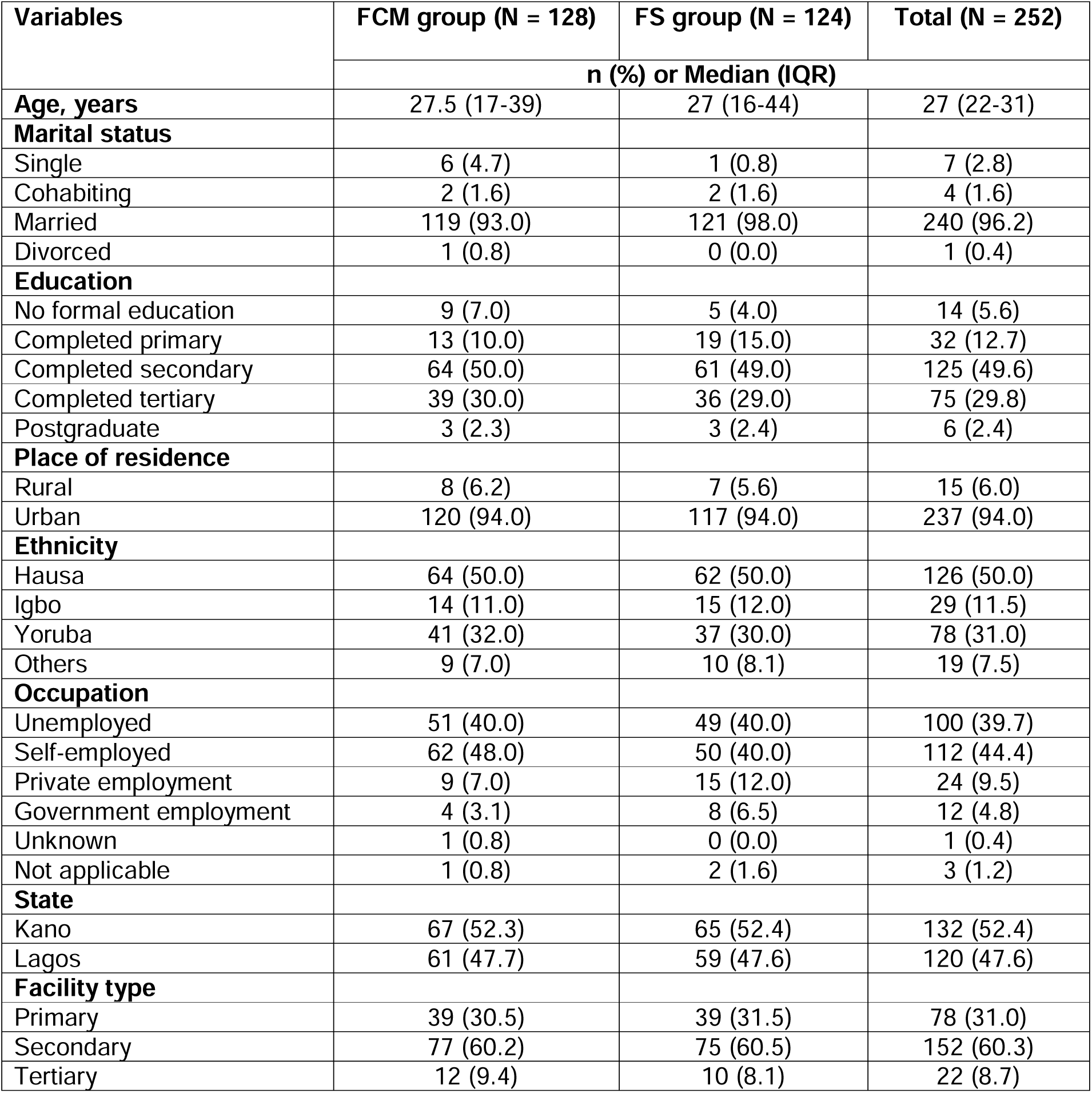
Sociodemographic characteristics of respondents.

| Variables | FCM group (N = 128) | FS group (N = 124) | Total (N = 252) |
| --- | --- | --- | --- |
|  | n (%) or Median (IQR) |  |  |
| **Age, years** | 27.5 (17-39) | 27 (16-44) | 27 (22-31) |
| **Marital status** |  |  |  |
| Single | 6 (4.7) | 1 (0.8) | 7 (2.8) |
| Cohabiting | 2 (1.6) | 2 (1.6) | 4 (1.6) |
| Married | 119 (93.0) | 121 (98.0) | 240 (96.2) |
| Divorced | 1 (0.8) | 0 (0.0) | 1 (0.4) |
| **Education** |  |  |  |
| No formal education | 9 (7.0) | 5 (4.0) | 14 (5.6) |
| Completed primary | 13 (10.0) | 19 (15.0) | 32 (12.7) |
| Completed secondary | 64 (50.0) | 61 (49.0) | 125 (49.6) |
| Completed tertiary | 39 (30.0) | 36 (29.0) | 75 (29.8) |
| Postgraduate | 3 (2.3) | 3 (2.4) | 6 (2.4) |
| **Place of residence** |  |  |  |
| Rural | 8 (6.2) | 7 (5.6) | 15 (6.0) |
| Urban | 120 (94.0) | 117 (94.0) | 237 (94.0) |
| **Ethnicity** |  |  |  |
| Hausa | 64 (50.0) | 62 (50.0) | 126 (50.0) |
| Igbo | 14 (11.0) | 15 (12.0) | 29 (11.5) |
| Yoruba | 41 (32.0) | 37 (30.0) | 78 (31.0) |
| Others | 9 (7.0) | 10 (8.1) | 19 (7.5) |
| **Occupation** |  |  |  |
| Unemployed | 51 (40.0) | 49 (40.0) | 100 (39.7) |
| Self-employed | 62 (48.0) | 50 (40.0) | 112 (44.4) |
| Private employment | 9 (7.0) | 15 (12.0) | 24 (9.5) |
| Government employment | 4 (3.1) | 8 (6.5) | 12 (4.8) |
| Unknown | 1 (0.8) | 0 (0.0) | 1 (0.4) |
| Not applicable | 1 (0.8) | 2 (1.6) | 3 (1.2) |
| **State** |  |  |  |
| Kano | 67 (52.3) | 65 (52.4) | 132 (52.4) |
| Lagos | 61 (47.7) | 59 (47.6) | 120 (47.6) |
| **Facility type** |  |  |  |
| Primary | 39 (30.5) | 39 (31.5) | 78 (31.0) |
| Secondary | 77 (60.2) | 75 (60.5) | 152 (60.3) |
| Tertiary | 12 (9.4) | 10 (8.1) | 22 (8.7) |

In comparing their perceptions of treatment, most women in the FCM and FS groups felt their providers explained treatment procedures “all the time” (85.9% and 75.8% respectively) and felt providers spoke in a language they could understand “all the time” (92.2% and 91.9% respectively). Most respondents also felt providers explained why they gave medicine “all the time” (90.6% and 92.7% respectively), felt they could ask any question “all the time” (75.6% and 73.4% respectively), and felt the providers took the best care of them “all the time” (91.4% and 93.6% respectively). Almost all the respondents (96.1% and 91.9% respectively) denied challenges with side effects. These perceptions were not statistically different among the two groups (p > 0.05).

A significantly higher proportion of respondents receiving FCM (82.8%) perceived their treatment to be “easy” compared to 63.7% receiving FS (p = 0.006) (Table 2). When asked about overall satisfaction with information and support received from providers, a significantly higher proportion of respondents in the FCM group (80.5%) rated the treatment as “excellent” compared to 62.1% in the FS group (p = 0.005). Significantly more people in the FCM vs the FS group (70.3 vs 50.0%) would also “very strongly recommend” the treatment to a family member (p=0.018) [Table 2].

**Table 2:** Comparison of perception of intravenous and oral iron treatment among respondents.

| Variables | Perception of FCM treatment (N = 128)<br>n (%) | Perception of FS treatment (N = 124)<br>n (%) | p-value |
| --- | --- | --- | --- |
| **Felt provider explained what had been done** |  |  | 0.122 |
| A few times | 2 (1.6) | 2 (2.4) |  |
| Most times | 16 (12.5) | 27 (21.8) |  |
| All the time | 110 (85.9) | 94 (75.8) |  |
| **Felt provider spoke in a language they could understand** |  |  | 0.941 |
| Most times | 10 (7.8) | 10 (8.1) |  |
| All the time | 118 (92.2) | 114 (91.9) |  |
| **Felt provider explained why he/she was giving medicine** |  |  | 0.889 |
| A few times | 3 (2.3) | 3 (2.4) |  |
| Most times | 8 (6.3) | 5 (4.0) |  |
| All the time | 116 (90.6) | 115 (92.7) |  |
| Missing | 1 (0.8) | 1 (0.8) |  |
| **Felt she could ask provider any question** |  |  | 0.850 |
| Never | 11 (8.7) | 9 (7.3) |  |
| A few times | 6 (4.7) | 6 (4.8) |  |
| Most times | 14 (11.0) | 18 (14.50) |  |
| All the time | 96 (75.6) | 91 (73.4) |  |
| **Felt providers took the best care of her they could** |  |  | 0.559 |
| A few times | 1 (0.8) | 0 (0.0) |  |
| Most times | 10 (7.8) | 8 (6.5) |  |
| All the time | 117 (91.4) | 116 (93.6) |  |
| **Ease of treatment administration** |  |  | **0.006** |
| Easy; no challenge | 106 (82.8) | 79 (63.7) |  |
| Little challenge | 19 (14.8) | 35 (28.2) |  |
| Moderate challenge | 3 (2.3) | 9 (7.3) |  |
| Lot of challenge | 0 (0.0) | 1 (0.8) |  |
| **Seriousness of side effects** |  |  | 0.164 |
| No challenge | 123 (96.1) | 114 (91.9) |  |
| Little challenge | 2 (2.3) | 9 (7.3) |  |
| Moderate challenge | 2 (1.6) | 1 (0.8) |  |
| **Overall satisfaction with information and support received** |  |  | **0.005** |
| Fair | 1 (0.8) | 1 (0.8) |  |
| Good | 24 (18.8) | 46 (37.1) |  |
| Excellent | 103 (80.5) | 77 (62.1) |  |
| **Would recommend treatment to a family member** |  |  | **0.018** |
| Very strongly do not recommend | 1 (0.8) | 2 (1.6) |  |
| Strongly do not recommend | 0 (0.0) | 2 (1.6) |  |
| Undecided | 1 (0.8) | 1 (0.8) |  |
| Recommend | 4 (3.1) | 12 (9.7) |  |
| Strongly recommend | 32 (25.0) | 45 (36.3) |  |
| Very strongly recommend | 90 (70.3) | 62 (50.0) |  |

**Table 3:**
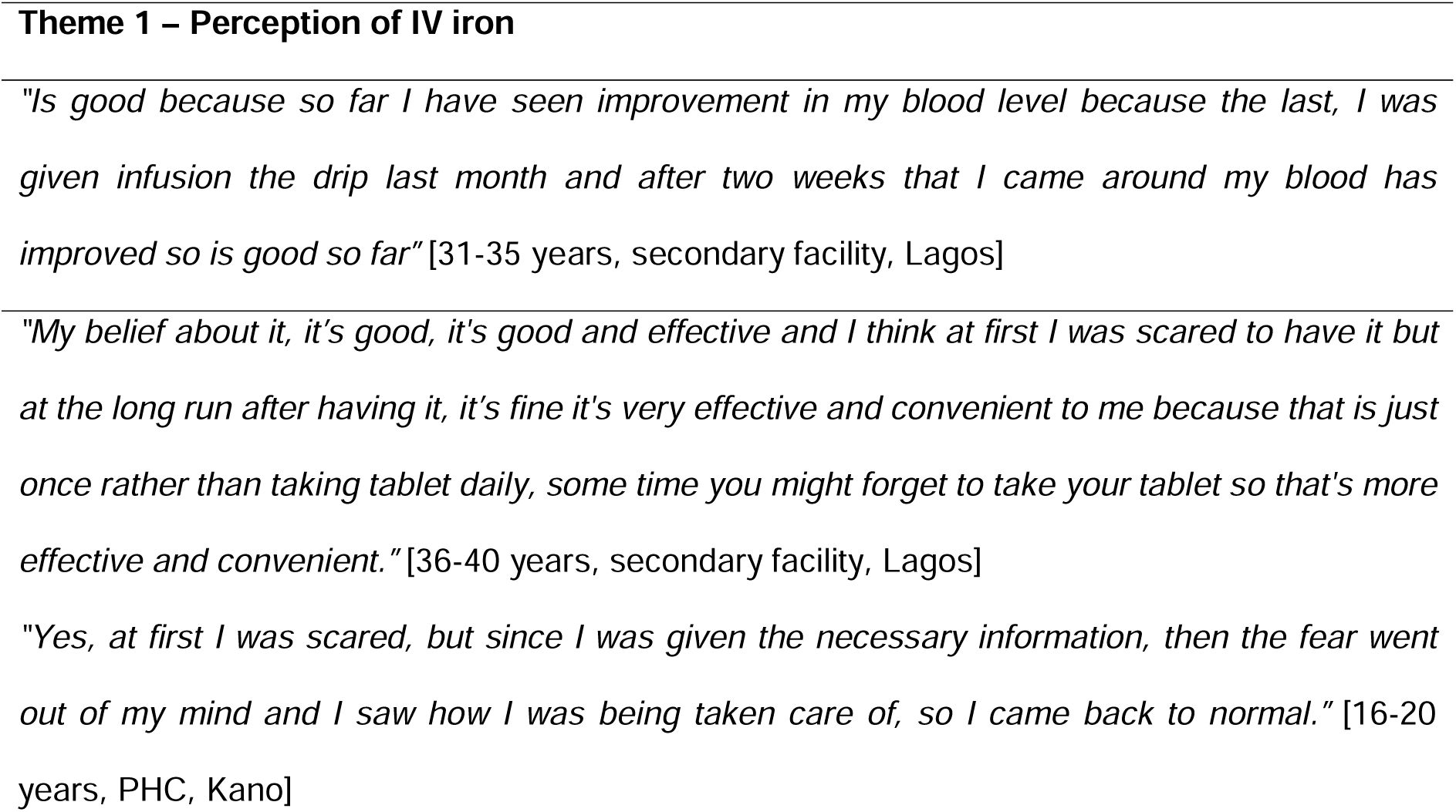
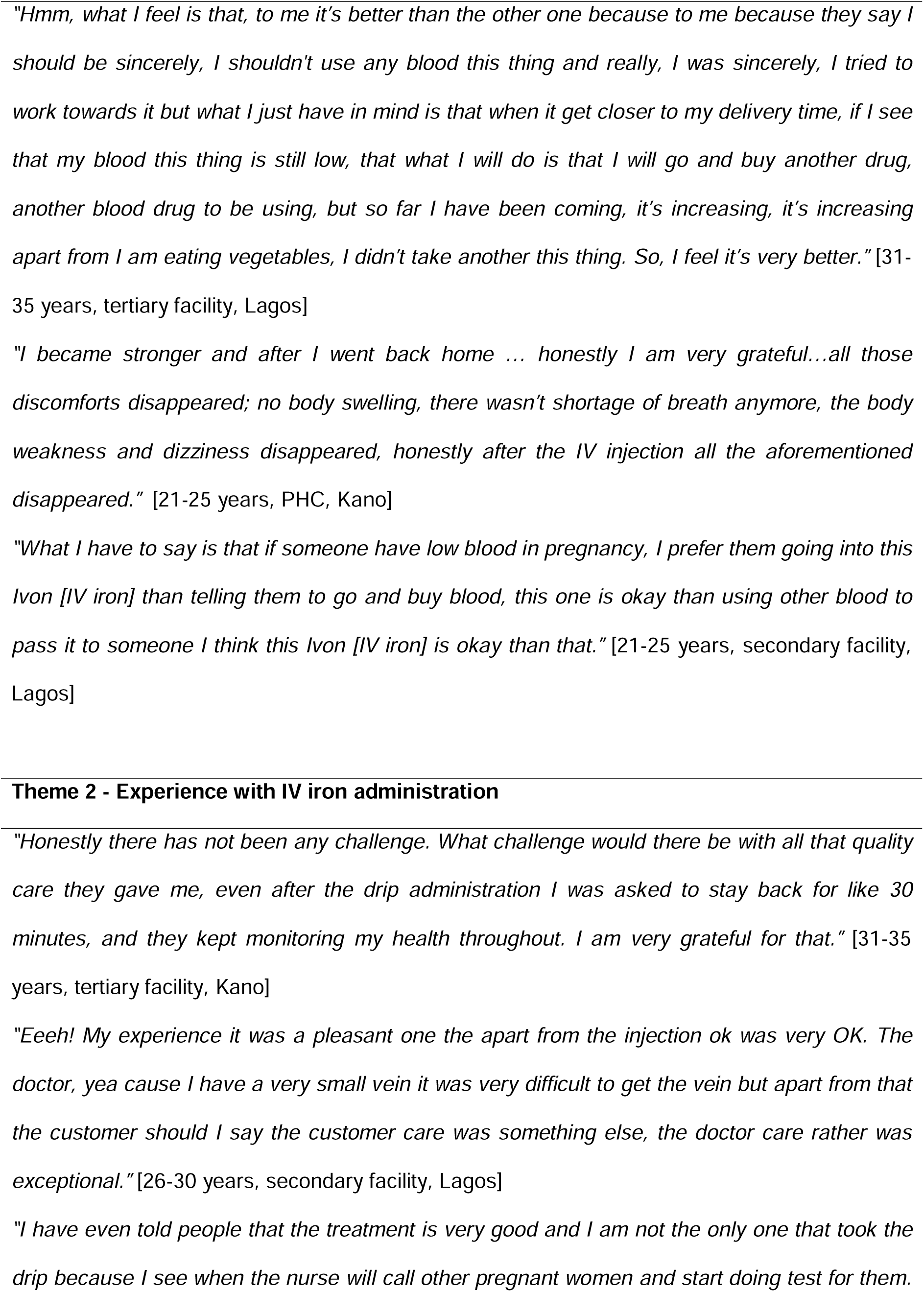
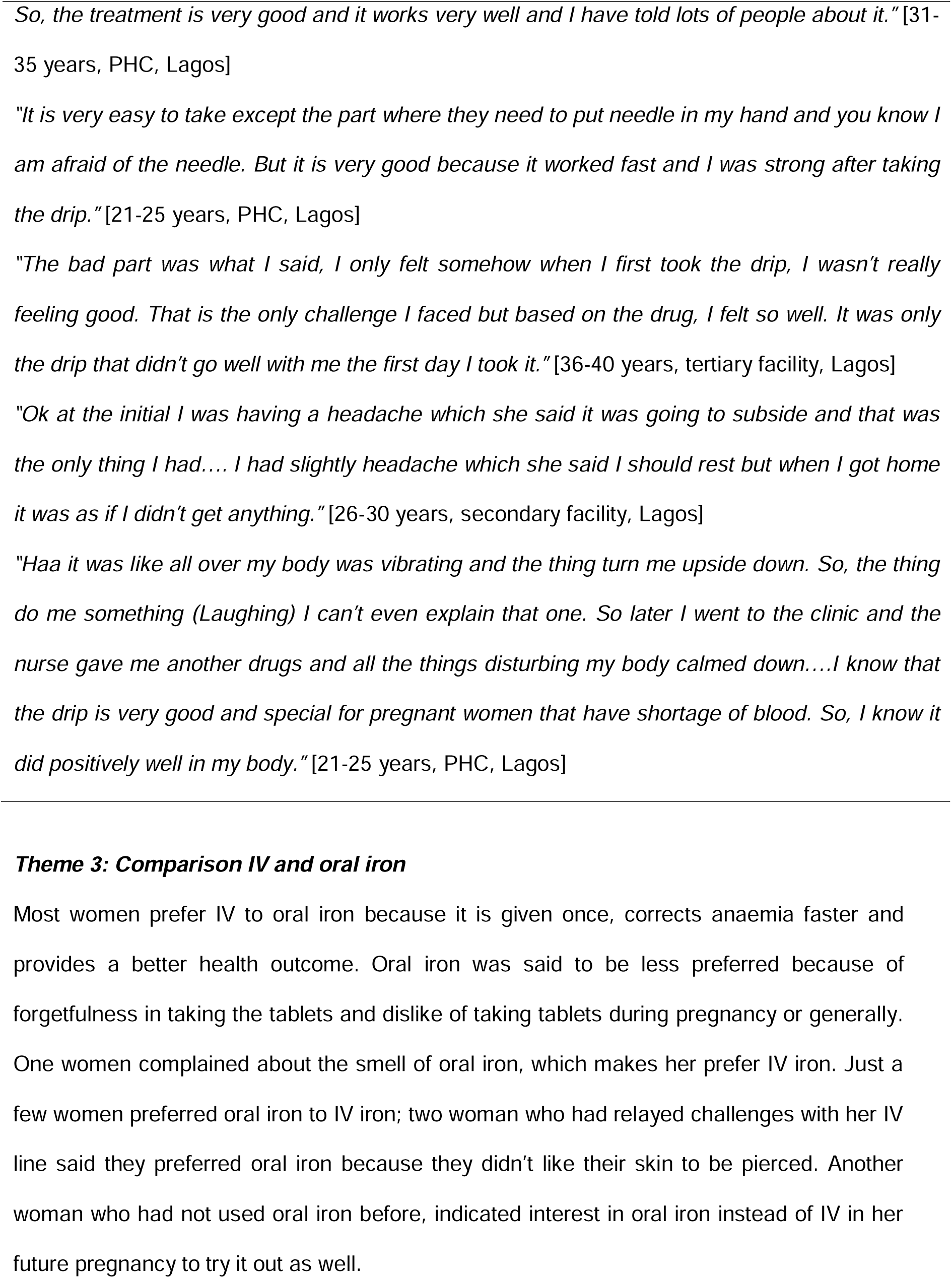
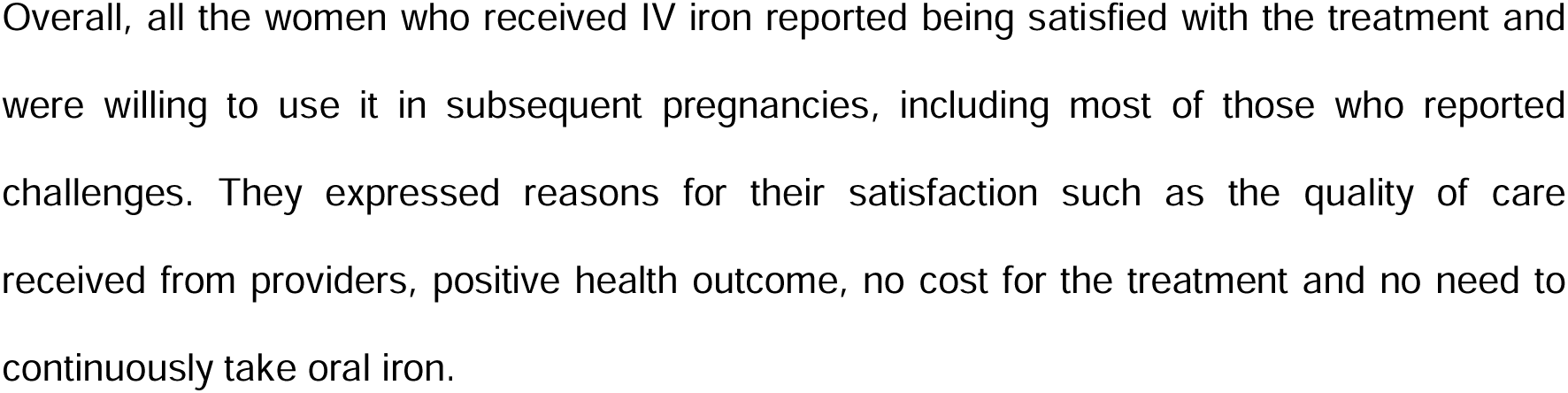
Illustrative quotes for themes 1 and 2.

**Table 4:**
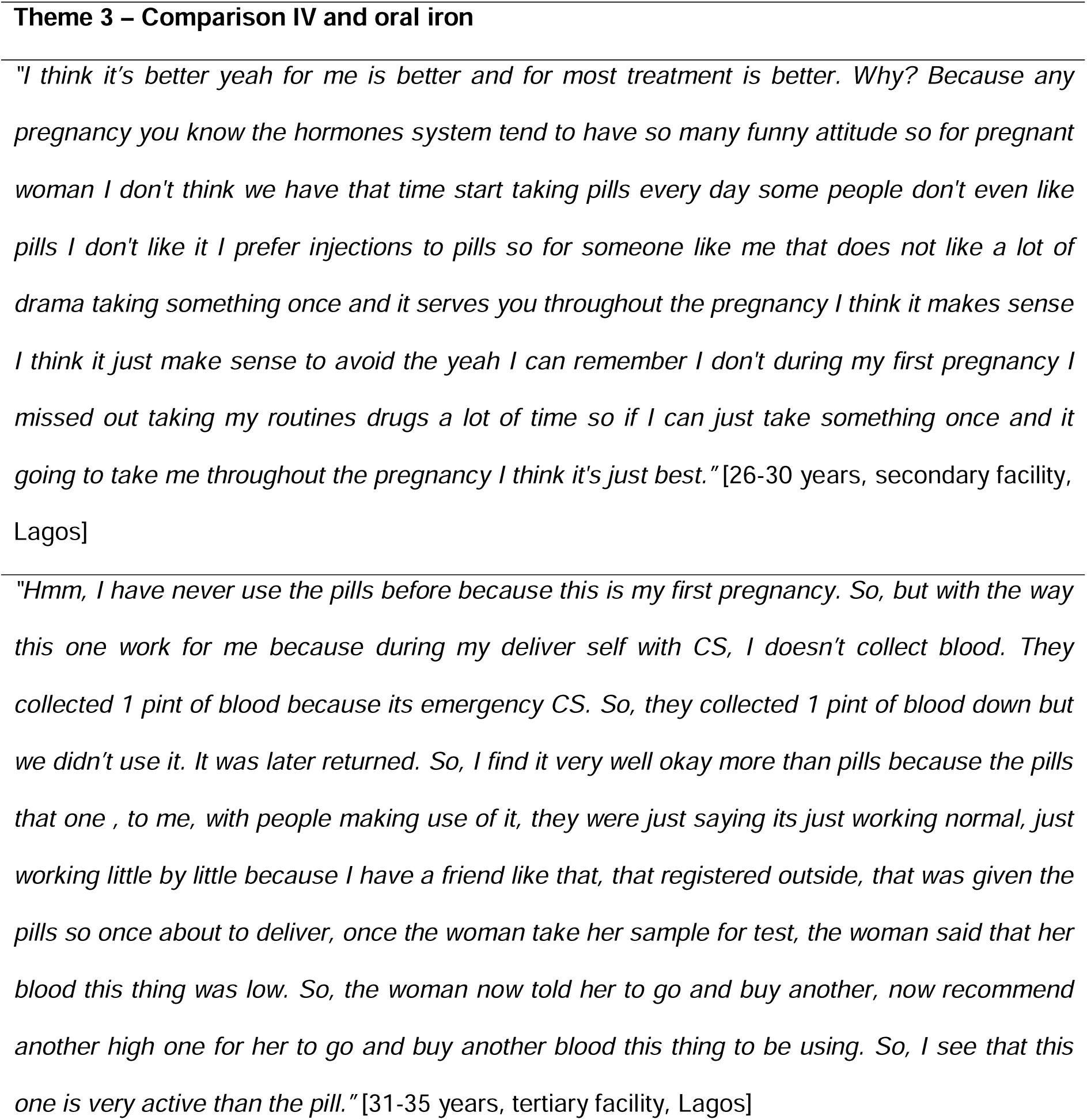
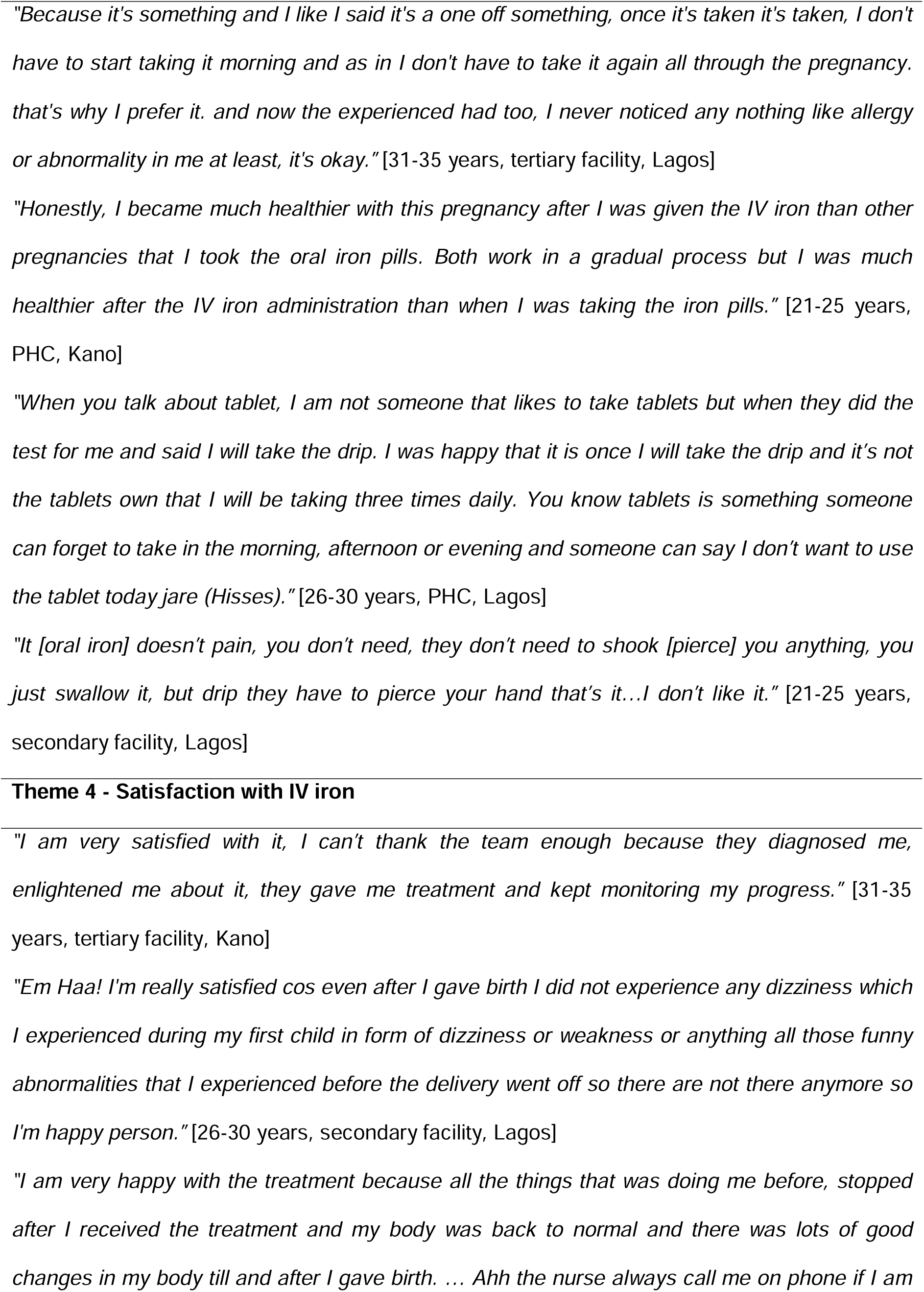
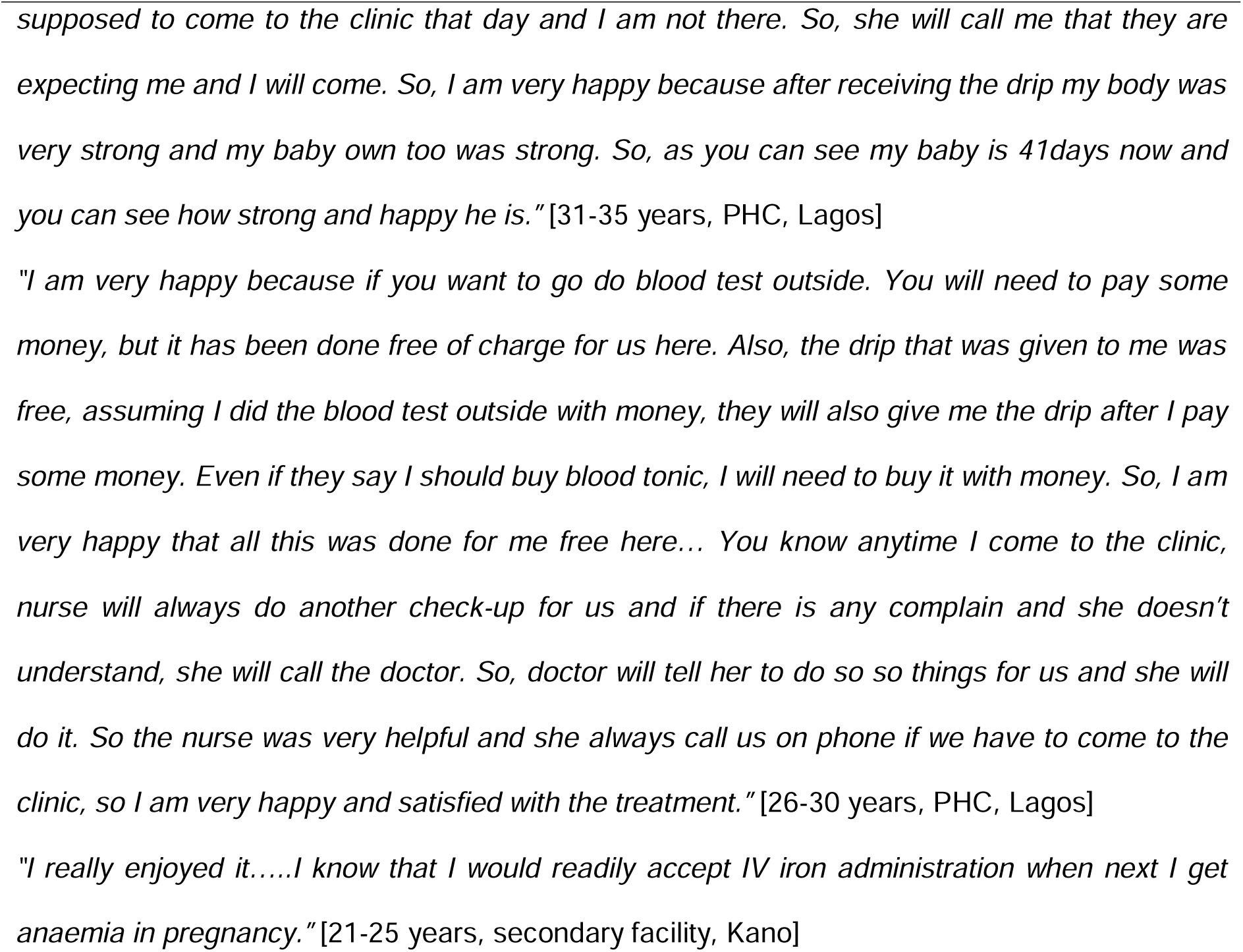
Illustrative quotes for themes 3 and 4.

### Qualitative results

Four main themes were identified: perception of IV iron, experience with IV iron administration, comparison of IV and oral iron, and satisfaction with IV iron therapy.

#### Theme 1: Perception of IV iron

Although some women were initially scared or doubtful of an unknown treatment, there was generally a positive perception about IV iron. It was perceived to be good, effective, convenient, and fast-acting. The information relayed by providers was perceived to allay fear or anxiety related to IV iron treatment. Some women reported immediate improvement in their anaemia symptoms after receiving IV iron. Several women noted that IV iron was advantageous as it reduced the need for blood transfusion.

#### Theme 2: Experience with IV iron administration

Most women reported having a positive experience with IV iron administration, highlighting the quality of care received from providers. Most reported not having side effects or other challenges and some women had already recommended the treatment to others. However, a few women reported challenges with setting the IV line and with side effects. The challenges with the IV line included repeated skin piercing, pain and injection phobia. Regarding the experience of side effects, one woman reported “not feeling good” the first day of the administration. Another woman reported having a transient headache, while two women reported having a fever the day after administration and one woman required administration of another drug to counter her side effects. Nevertheless, the women who reported challenges felt the treatment worked well for them.

#### Theme 3: Comparison IV and oral iron

Most women prefer IV to oral iron because it is given once, corrects anaemia faster and provides a better health outcome. Oral iron was said to be less preferred because of forgetfulness in taking the tablets and dislike of taking tablets during pregnancy or generally. One women complained about the smell of oral iron, which makes her prefer IV iron. Just a few women preferred oral iron to IV iron; two woman who had relayed challenges with her IV line said they preferred oral iron because they didn’t like their skin to be pierced. Another woman who had not used oral iron before, indicated interest in oral iron instead of IV in her future pregnancy to try it out as well.

#### Theme 4: Satisfaction with IV iron

Overall, all the women who received IV iron reported being satisfied with the treatment and were willing to use it in subsequent pregnancies, including most of those who reported challenges. They expressed reasons for their satisfaction such as the quality of care received from providers, positive health outcome, no cost for the treatment and no need to continuously take oral iron.

### Data integration

Integration of key quantitative and qualitative results showed high convergence as illustrated in Table 5.

**Table 5:**
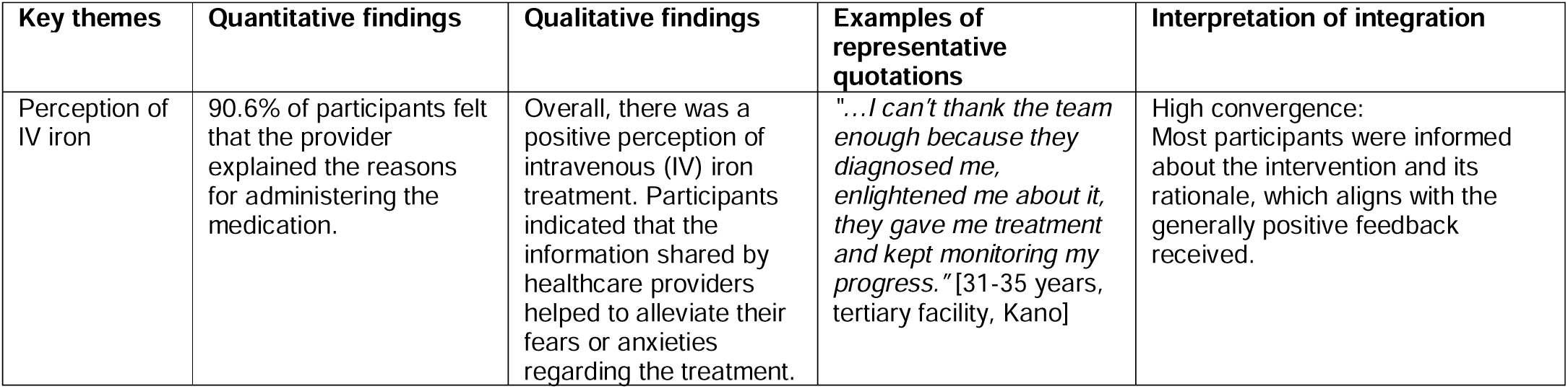
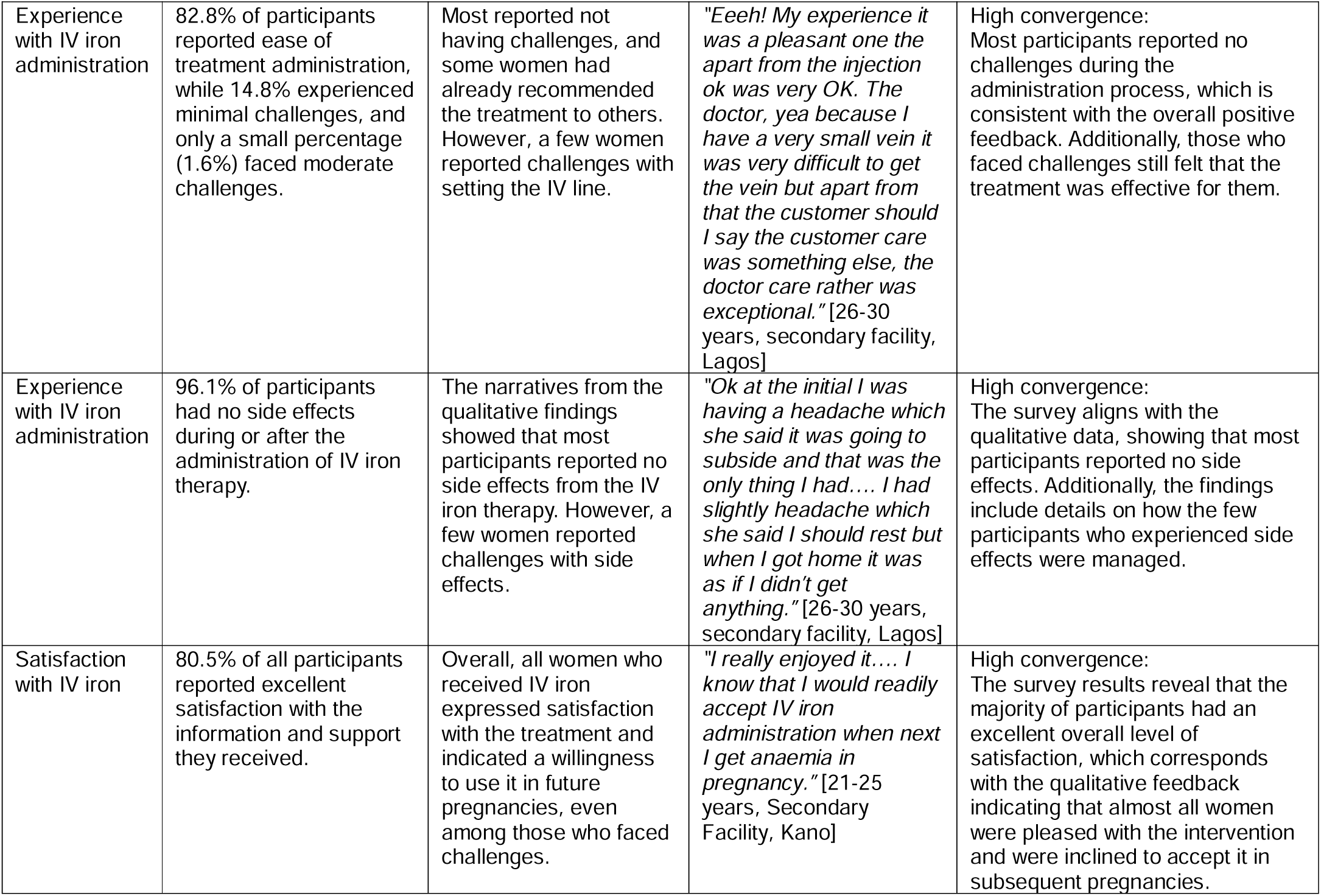
illustrating the integration of quantitative and qualitative findings.

## Discussion

This study evaluated the perceptions, experiences, and satisfaction of pregnant women receiving intravenous iron (ferric carboxymaltose; FCM) versus oral iron (ferrous sulphate; FS) for the treatment of anaemia in pregnancy within a Type 1 hybrid effectiveness-implementation framework. By employing a concurrent mixed-methods design, we were able to integrate quantitative survey data with qualitative in-depth interviews, thereby providing a comprehensive understanding of patient experiences that are crucial for guiding the future scale-up of IV iron in routine antenatal care.

The survey findings reveal a significantly higher proportion of women in the IV iron group who rated treatment as’excellent’ (80.5%). Also, many participants highlighted the convenience of a single-dose administration and the elimination of the daily pill burden—a factor that not only improved adherence but also alleviated common concerns associated with oral iron therapy. For instance, one respondent remarked, “Taking something once and knowing it will last throughout pregnancy is the best option for me,” which mirrors the favorable trends seen in the survey responses.

A significant theme emerging from the qualitative data highlighted the pivotal role of healthcare providers in fostering positive perceptions of treatment. Participants frequently noted that clear and empathetic communication played a key role in alleviating initial concerns about IV administration. This effective patient-provider interaction, supported by high ratings for provider explanations and qualitative feedback, emphasizes the importance of personalised patient education and reciprocal communication in promoting treatment acceptance. These findings align with earlier studies that stress the influence of strong communication on enhancing adherence and patient satisfaction [24, 25].

From an implementation science standpoint, the high levels of acceptability and patient satisfaction linked to IV iron indicate that FCM could be a practical alternative to oral iron, particularly in regions with high prevalence of anaemia. However, it is important to acknowledge that our study took place in a controlled trial setting, where patients received free treatment and were closely monitored. These conditions might have shaped patient perceptions positively and may not fully reflect real-world healthcare scenarios. While the findings are encouraging, large-scale implementation of IV iron treatment for AIP would require addressing logistical barriers, such as cost and health system readiness. While participants in the IVON trial received treatment at no cost, affordability remains a major determinant of sustainability in routine antenatal care.

### Strengths and limitations

A key strength of this study is its robust mixed-methods design, which integrates quantitative survey data with IDIs to provide a comprehensive view of client satisfaction with FCM versus FS. The inclusion of qualitative interviews allowed us to extract rich, firsthand comments on treatment experiences and satisfaction levels. This dual-method approach not only enhances the credibility of our findings but also offers deeper insights into the factors driving patient preference and satisfaction. This study is also the first to assess the perceptions and experiences of FCM among Nigerian women with the added strength of comparisons to those of women receiving the oral iron standard of care.

However, there are limitations to consider. First, while the qualitative interviews added valuable context, they were conducted within a controlled trial setting, which may have influenced the participants’ positive perceptions compared to a routine clinical environment. Second, the potential for recall bias exists due to the time lag between treatment administration and the exit interviews. Finally, despite the purposive sampling strategy for the qualitative phase aimed at capturing a range of experiences, the sample may not fully represent the diverse perspectives of all pregnant women receiving FCM and FS in Nigeria.

## Conclusion

Positive patient perception about FCM, positions it as a preferred alternative to traditional oral iron therapy among pregnant women who participated in the IVON trial, offering clear advantages in the terms of the ease of use and overall patient satisfaction. These findings not only support the clinical benefits of FCM but also provide compelling evidence for its potential implementation in routine antenatal care. To capitalize on these benefits, policymakers and healthcare providers must address systemic barriers—including cost, logistical challenges, and the need for comprehensive provider training—to facilitate the broader adoption of IV iron therapy. Future research should focus on real-world implementation studies, including cost-effectiveness analyses and long-term follow-up assessments that evaluate the durability of treatment effects—not only in terms of clinical outcomes such as sustained improvements in haemoglobin levels and reduction of anaemia-related complications, but also regarding social impacts, including patient satisfaction, quality of life, and broader health system integration—to further validate the integration of IV iron into standard maternal healthcare practices.

## Data Availability

Data cannot be shared publicly because of concerns of breach of confidentiality. Data are available from the Center of Clinical Trials, Research and Implementation Science Institutional Data Access (contact via) for researchers who meet the criteria for access to confidential data.

## Acknowledgements

We wish to acknowledge the IVON trial study team in Lagos and Kano states for their support for this study. We appreciate the research assistants (Binta Umar Abdullahi, Amina Abubakar, Rukayya Sadiqa Isa, Judith Amos, Jennifer Ejiofor, Oluchi Ozonu, Jacob Igologba, Kehinde Adeshina, Donald Ezinwanne) for the hard work in collecting data and the postpartum women who took time to provide the information required for this study.

## Supporting information

**S1 Checklist.** Standards for Reporting Implementation Studies (StaRI) checklist

